# Genome Sequencing Identifies Monogenic Causes in Adults with Metabolic Diseases

**DOI:** 10.1101/2025.11.14.25334651

**Authors:** Volkan Okur, Amanda Halstrom, John N. Falcone, Maurice A. Hurd, Sarah L. Stewart, Katerine Claudio, Jyothi Manohar, Fana Dealla, Sonal Kumar, Michele Yeung, Gregory Dakin, Omar Bellorin-Marin, Cheguevara Afaneh, Lisa C. Hudgins, Esther Wei, Laura Gingras, Alexandra King, Judy Tung, Atteeq U. Rehman, Amanda Thomas-Wilson, Saurav Guha, Avinash Abhyankar, Ashley L. Wilson, Shahid Yar Khan, Sowmya Thirumalai Srinivasa, Shruti Phadke, Priya Krithivasan, Caroline Nava, Shuibing Chen, Ryan Smith, Theresa Y. MacDonald, Megan J. Ritter, Laura C. Alonso, Olivier Elemento, Miriam S. Udler, Jessica M. Peña, Vaidehi Jobanputra, Marcus D. Goncalves

## Abstract

**Purpose:** A subset of metabolic diseases is caused by rare monogenic variants. Next generation sequencing (NGS)-based testing offers a promising approach for identifying such variants. However, its application in clinical diagnostics for metabolic diseases is limited, and the diagnostic yield is unknown.

**Methods:** We performed clinical genome sequencing (GS) on 560 adults seen in New York clinical practices between August 2020 and December 2023. Participants presented with hyperlipidemia/hypertriglyceridemia (HLD/HTG), pre-diabetes, Type 2 diabetes mellitus (T2DM), and metabolic dysfunction-associated fatty liver disease/steatohepatitis (MAFLD/MASH). Variants in a curated set of 90 genes associated with monogenic forms of these conditions were classified as Pathogenic (P), Likely Pathogenic (LP), and Variant of Uncertain Significance (VUS) using ACMG and ClinGen Sequence Variant Interpretation Working Group guidelines. P/LP variants in ACMG secondary findings (v3.1) genes were also reported with participant consent.

**Results:** The cohort included a female-to-male ratio of 1.7, with 18.6% African American and 22.6% Latino participants. The most common primary enrollment diagnoses were HLD/HTG (25%), T2DM (9%), pre-diabetes (7%), and MAFLD/MASH (4%). Many participants had multiple conditions (42% with two, 12% with three).

Approximately one-third had reportable variants with 6% classified as P/LP. The most common P/LP variants were in APOB and LDLR.

**Conclusions:** The prevalence of clinically significant (P/LP) variants related to primary metabolic disease in this cohort was 6%. An additional 5.5% of participants had P/LP variants in genes recommended for return as ACMG secondary findings. Future studies should focus on refining participant selection for genome sequencing to optimize its diagnostic and clinical value.

## INTRODUCTION

Metabolic diseases such as diabetes, obesity, hyperlipidemia/hypertriglyceridemia (HLD/HTG), and metabolic dysfunction-associated fatty liver disease/steatohepatitis (MAFLD/MASH) represent a significant public health risk worldwide, particularly in developed countries.^1^ While environmental factors such as diet and exercise play critical roles in the etiology of these conditions, familial studies have demonstrated for decades that genetic factors also contribute to their development.^2^ Genomic variants can contribute to the development of metabolic diseases via polygenic and monogenic mechanisms.

In monogenic conditions, the majority of risk can be attributed to pathogenic variants in a single gene, either in a monoallelic or biallelic state. For instance, causal variants in genes encoding for the low density lipoprotein receptor (LDLR), apolipoprotein B-100 (APOB), and proprotein convertase subtilisin/kexin type 9 (PCSK9) result in familial hypercholesterolemia, a condition characterized by elevated plasma low-density lipoprotein cholesterol (LDL) levels and premature cardiovascular disease.^3^ Similarly, monogenic forms of diabetes such as maturity-onset diabetes of the young (MODY), involve specific genes (e.g. GCK, HNF1A, HNF1B, HNF4A) that cause beta-cell dysfunction, resulting in hyperglycemia.^4^ Monogenic and syndromic obesity arise from variants in genes related to regulating appetite and food intake in the central nervous system as well as in adipocyte function (BBS, LEP, LEPR, MC4R, POMC, SIM1).^5^ Congenital forms of lipodystrophy result from loss-of-function variants in genes associated with adipocyte development or structure (AGPAT2, CAV1, LMNA, PPARG), leading to partial or complete loss of adipose depots.^6^

Next generation sequencing (NGS)-based technologies, including targeted gene panels (TGP), exome (ES), and genome sequencing (GS), are extensively utilized to obtain molecular diagnoses in individuals with various health conditions such as neurodevelopmental disorders, ophthalmologic disorders, cancer, and congenital endocrine disorders.^7,8^ The molecular diagnoses not only establish a definitive diagnosis but also inform disease management, family planning, and cascade screening. Genome-wide association studies (GWAS) have frequently been utilized in identifying multiple genes and variants contributing to the pathogenesis of multifactorial metabolic diseases.^9^ It is well-established that metabolic diseases such as diabetes, obesity, and hyperlipidemia can sometimes result from rare, high-impact pathogenic variants in genes associated with monogenic forms of these diseases. However, pinpointing a specific monogenic cause in adults presenting with metabolic diseases remains challenging due to non-specific biochemical and physical findings across multiple gene-related conditions. While NGS technologies allow for comprehensive investigation of monogenic causes of metabolic diseases, this approach has not yet gained widespread acceptance in metabolic disease clinics apart from familial hypercholesterolemia cohorts.^10–12^ Consequently, the diagnostic yield of GS in individuals with multiple common and overlapping metabolic diseases remains unknown.

## METHODS

This study was approved by the Weill Cornell Medicine Institutional Review Board, and all participants provided written informed consent for clinical GS test.

### Study Population

Between August 2020 and December 2023, 788 eligible individuals were offered participation in the study and 560 agreed to participate. The primary reasons for non-participation were concerns regarding the security of personal genetic information and a perceived lack of personal benefit from study participation. Two participants were withdrawn after enrollment due to delays in blood sample acquisition and excessive time burden placed on participants. Participants aged 18 years or older who met one or more inclusion criteria were recruited from specific Weill Cornell Medicine clinics: Internal Medicine, Endocrinology and Weight Management, Bariatric Surgery, Preventive Cardiology, Lipid Control Center, and Hepatology. Enrollment criteria were: Pre-diabetes defined as hemoglobin A1C between 5.7-6.4%, fasting plasma glucose 100 and 125 mg/dL, or 2h plasma glucose during 75g oral glucose tolerance test between 140 and 199 mg/dL; Type 2 diabetes (T2DM) defined as hemoglobin A1C ≥6.5%, fasting plasma glucose ≥126 mg/dL, random plasma glucose ≥200 mg/dL, or 2h plasma glucose during 75g oral glucose tolerance test ≥200 mg/dL; HLD/HTG defined by at least one of the following: Total cholesterol >200 mg/dL, low-density lipoprotein (LDL) cholesterol >130 mg/dL or on pharmacologic agents with explicit purpose to decrease LDL, high-density lipoprotein (HDL) cholesterol <40 mg/dL for males and <50 mg/dL for females, triglycerides >250 mg/dL or on pharmacologic agents with explicit purpose to decrease triglycerides; high risk for MAFLD with BMI ≥25 kg/m^2^ and ALT >25 IU/L in females and >33 IU/L in males; and MASH defined as: Hepatic steatosis by imaging or histology, Baseline Fibroscan CAP score > 238 dB/m, ALT >25 IU/mL in females and >33 IU/mL in males with no compelling etiology for hepatic steatosis. Exclusion Criteria were age <18 years, diagnosis of compensated or decompensated cirrhosis, and known or suspected hepatocellular carcinoma.

### Gene Panel Selection

A total of 90 genes were selected for their association with metabolic disease traits, including non-syndromic (e.g. LDLR) and syndromic (e.g. ALMS1), as detailed in Supplementary Table 1. Additionally, genes recommended by the American College of Medical Genetics as secondary findings gene list were included provided the participant consented to receive results for these findings (versions v2 or v3.1) at the time of testing (Supplementary Table 1).^13,14^ Notably, LDLR, APOB, PCSK9, HNF1A, and LMNA from the metabolic disease gene panel also overlap with the ACMG secondary findings gene lists.

### Genome Sequencing

Peripheral whole blood samples were collected in EDTA tubes in each participating clinic and stored at 4 °C before weekly shipment to the New York Genome Center molecular diagnostic laboratory. Proband only clinical GS test was performed.

DNA extraction was performed using standard methods. GS libraries were prepared using either KAPA Hyper Prep kit (KAPA Biosystems) or TruSeq DNA Nano/TruSeq DNA PCR-free library preparation kit (Illumina, CA). Sequencing was performed on Illumina HiSeq X or NovaSeq 6000 instruments (Illumina, CA) with 150-bp paired-end reads. Human genome builds 37 (GRCh37/hg19) or 38 (GRCh38/hg38) were used as reference sequence assemblies during the early and late stages of the study, respectively. All genomes achieved at least 30X±3X mean sequence coverage, with a minimum of 85% of bases were sequenced to at least 20X coverage.

### Variant Analysis

GS data of each participant was analyzed for the variants in the 90 genes included in the metabolic diseases panel. Single nucleotide (SNV) and copy number (CNV) variants were classified as variant of uncertain significance (VUS), likely pathogenic (LP), or pathogenic (P) based on American College of Medical Genetics and Genomics/Association of Molecular Pathology guidelines^15^ and modifications recommended by ClinGen’s Sequence Variant Interpretation working group (https://clinicalgenome.org/working-groups/sequence-variant-interpretation). Reportable variants associated with the phenotype or the primary indication for testing and in ACMG secondary findings genes were confirmed using an orthogonal method such as dideoxy terminator sequencing (Sanger sequencing) or genome-wide SNP microarray. These confirmed variants were reported with detailed gene and variant curations. Variants not clearly relevant to the testing indication, such as VUS in a gene on the 90-gene panel but associated with a different phenotype or a single P/LP variant in a recessive disease gene regardless of phenotype match, were reported in a supplementary section. This section included summary-level details such as gene, associated disease, variant nomenclature, and classification. This supplementary reporting aimed to enable reverse phenotyping. For ACMG secondary findings, only P/LP variants were reported.

### Ancestry Analysis

In addition to collecting self-reported race and ethnicity, we also performed genomic ancestry analysis using the fastNGSadmix software to estimate admixture proportions, and the ‘prcomp’ function from the ‘stats’ package in R to perform Principal Component Analysis.

## RESULTS

Demographic characteristics of the study population are summarized in Table 1. The mean age was 55 ± 16, with a median age of 57 years (range: 20-93). The female-to-male ratio was 1.7:1, with no significant difference in median age distribution based on sex assigned at birth. The majority of participants self-reported as White/Non-Latino and resided in New York City or the surrounding tri-state area (97%). Twenty-seven participants (5%) declined to disclose their race and ethnicity. Genomic ancestry analysis was largely consistent with self-reported race/ethnicity. However, mixed racial components were predominant among participants who identified themselves as ‘other’ in the race category (Supplementary Figure 1).

**Table 1.** Demographic characteristics of the study population.

|  |  |
| --- | --- |
| Sex, n (%) |  |
| Female | 353, 63% |
| Male | 207, 37% |
| Total | 560, 100% |
| Age, mean [SD] | 55 [± 16] |
| Age, median [range] | 57 [20-93] |
| Race, self-reported, n (%) |  |
| Asian | 65, 11.6% |
| Black or African American | 104, 18.6% |
| White | 261, 46.6% |
| Other | 98, 17.5% |
| Declined/Unknown | 32, 5.7% |
| Ethnicity, n (%) |  |
| Hispanic or Latino | 126, 22.6% |
| Non-Hispanic or Latino | 418, 74.6% |
| Declined/Unknown | 16, 2.8% |
| State of Residence |  |
| New York | 509, 90.8% |
| New Jersey | 34, 6.1% |
| Connecticut | 10, 1.8% |
| Other | 7, 1.3% |
| Primary Indications* |  |
| HLD/HTG | 424, 76% |
| T2DM | 229, 41% |
| Pre-diabetes | 134, 24% |
| MASH/MAFLD | 134, 24% |
| Two indications | 235, 42% |
| Three indications | 69, 12% |
| Comorbidities |  |
| BMI |  |
| ≥ 40 | 74, 13% |
| [35-40) | 79, 14% |
| [30-35) | 110, 20% |
| [25-30) | 153, 27% |
| [18-25) | 129, 23% |
| < 18 | 4, <1% |
| CAD | 210, 38% |
| Hypertension | 35, 6% |

\* This distribution does not indicate any single patient's presenting complaint but the presence of each of four primary indications across all patients. Distribution of primary indications as presenting phenotypes is provided in Figure 1.
Abbreviations: BMI; Body mass index, CAD; Coronary artery disease, HLD/HTG; Hyperlipidemia/Hypertriglyceridemia, MASH/MAFLD; Metabolic dysfunction-associated fatty liver disease/steatohepatitis, SD; standard deviation, T2DM; Type 2 Diabetes Mellitus

The most common primary enrollment diagnoses were HLD with or without hypertriglyceridemia (HTG) (n = 140, 25%), T2DM (n = 51, 9%), Pre-diabetes (n = 38, 7%), and MAFLD or MASH (n = 25, 4%). A substantial number of participants had more than one metabolic disease consistent with the concept of metabolic syndrome (Figure 1). Other comorbidities like obesity/overweight (n = 416, 74%), hypertension (n = 210, 38%), and coronary artery disease (n = 35, 6%) were also common in this population (Table 1).

**Figure 1.**
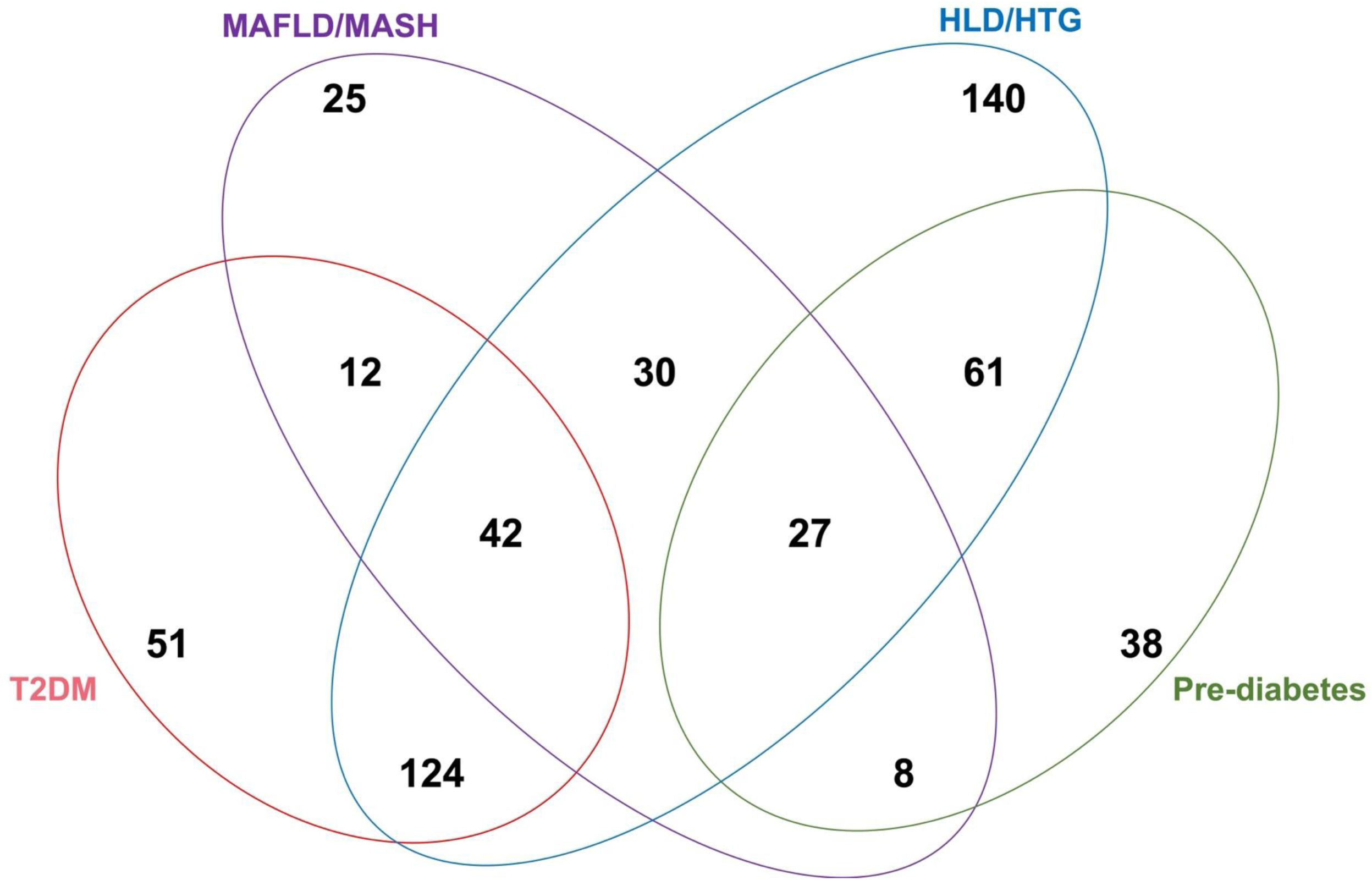
A four-way Venn diagram of the primary enrollment indications. HLD/HTG; hyperlipidemia/hypertriglyceridemia, MAFLD/MASH; metabolic dysfunction-associated fatty liver disease/steatohepatitis, T2DM; type 2 diabetes mellitus.

A total of 223 variants were reported in 175 participants (31%). These findings were relevant to the reported phenotype (Figure 2). Of the 223 reported variants, 33 P/LP variants (found in 31 participants, 6%) were directly associated with the phenotype. HLD/HTG either isolated (n = 18) or in combination with other phenotypes (n = 8) was the most common indication among participants with P/LP variants. The most frequently detected variants were in *LDLR* (*n* = 10), *APOB* (*n* = 6), and *LPL* (*n* = 3). Additional variants included *PPARG*, *APOE*, and *GNAS* (*n* = 2 in each), *NR0B2*, *GCK*, *LMNA*, *SIM1*, *MC4R*, *HNF1B*, *ABCG5*, *APOA5*, *ABCA1*, *GPIHBP1* (*n* = 1 in each). The variant in *APOA5* was a ∼165 Kb deletion encompassing the entire gene and all the other reported variants were single nucleotide variants (SNVs). Notably, one participant was homozygous for a pathogenic *LDLR* variant, and another participant carried two pathogenic variants in *LDLR*. All remaining participants were heterozygous for the detected variants (Table 2).

**Figure 2.**
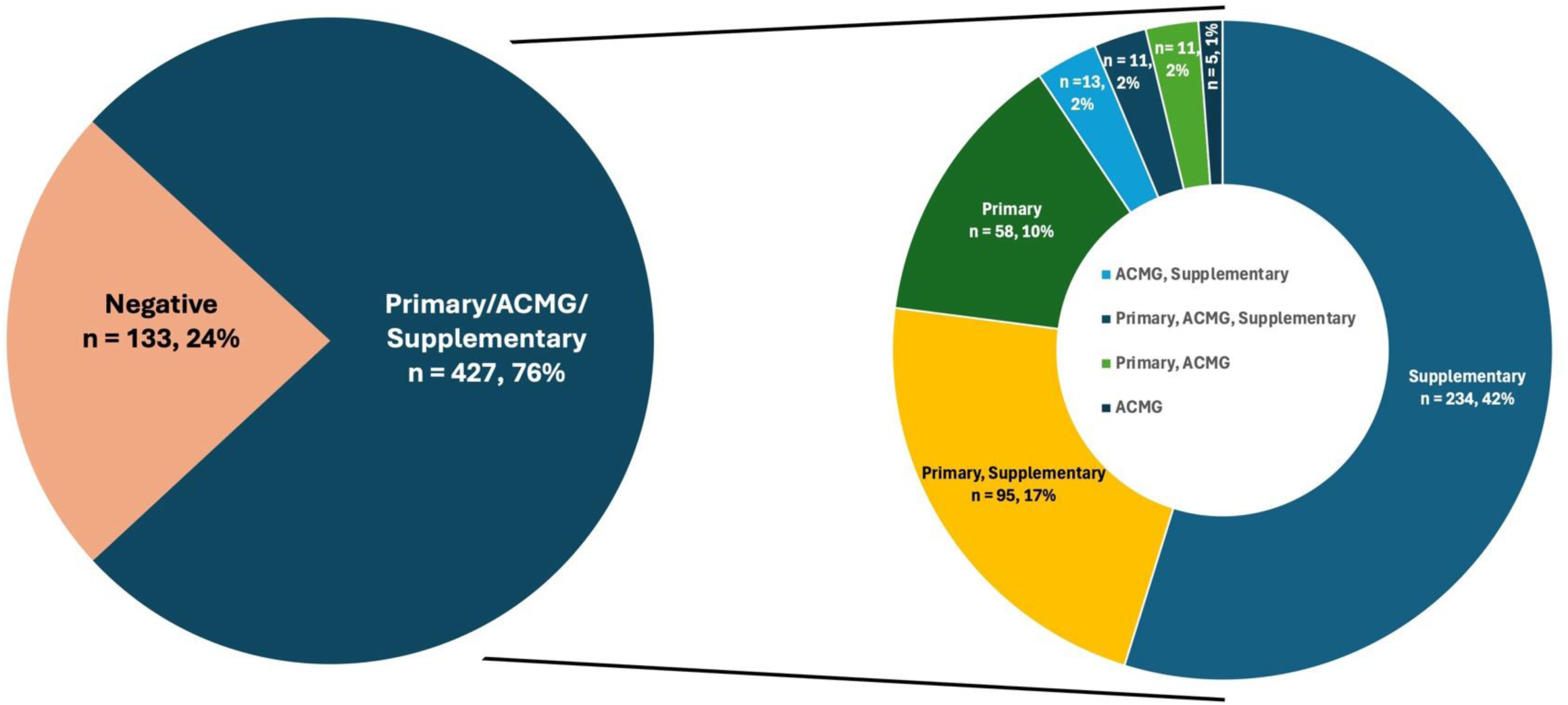
Distribution of participants receiving variants by report section. Left: distribution of entire cohort. Right: distribution of report sections in participants receiving at least one variant reported.

**Table 2.** Pathogenic/Likely pathogenic variants reported in participants with metabolic diseases.

| Participant ID | Sex | Age range | Indication | Gene (Transcript) | Variant | Classification | Disease | Results explain the indication |
| --- | --- | --- | --- | --- | --- | --- | --- | --- |
| 1 | M | 51-55 | HLD/HTG | LDLR<sup>b</sup><br>(NM_000527.4) | c.654_656del p.(Gly219del) | P | Hypercholesterolemia, familial, 1 [AD/AR; MIM#143890] \| LDL cholesterol level QTL2 [AD/AR; MIM#143890] | Yes |
| 2 | M | 21-25 | HLD/HTG |  | c.1055G>A p.(Cys352Tyr)<sup>a</sup> | LP |  | Yes |
| 3 | M | 46-50 | HLD/HTG |  | c.253C>T p.(Gln85Ter) | P |  | Yes |
| 4 | M | 21-25 | HLD/HTG |  | c.654_656del p.(Gly219del) | P |  | Yes |
| 5 | F | 41-45 | HLD/HTG |  | c.1A>T p.(Met1) | P |  | Yes |
| 5 | F | 41-45 | HLD/HTG |  | c.418G>A p.(Glu140Lys) | P |  | Yes |
| 6 | F | 26-30 | HLD/HTG |  | c.103C>T p.(Gln35Ter) | P |  | Yes |
| 7 | F | 56-60 | HLD/HTG |  | c.1618G>A p.(Ala540Thr) | LP |  | Yes |
| 8 | F | 56-60 | HLD/HTG |  | c.782G>T p.(Cys261Phe) | P |  | Yes |
| 9 | F | 41-45 | HLD/HTG |  | c.1195G>A p.(Ala399Thr) | LP |  | Yes |
| 10 | M | 56-60 | HLD/HTG+<br>MAFLD/MA<br>SH | APOB<sup>b</sup><br>(NM_000384.2) | c.6673dup<br>p.(Thr2225AsnfsTer4) | P | Hypobetalipoproteinemia [AR; MIM#615558] \|<br>Hypercholesterolemia, familial, 2 [AD; MIM#144010] | Yes |
| 11 | M | 31-35 | HLD/HTG+P<br>re-Diabetes |  | c.10579C>T p.(Arg3527Trp) | P |  | Yes |
| 12 | F | 51-55 | T2DM+MAFLD/MASH |  | c.6034C>T p.(Arg2012Ter) | LP |  | Yes, partially |
| 13 | F | 66-70 | HLD/HTG |  | c.10580G>A p.(Arg3527Gln) | P |  | Yes |
| 14 | F | 51-55 | HLD/HTG |  | c.10579C>T p.(Arg3527Trp) | P |  | Yes |
| 15 | F | 36-40 | T2DM+MAFLD/MASH |  | c.6034C>T p.(Arg2012Ter) | P |  | Yes |
| 16 | M | 66-70 | T2DM | LPL<br>(NM_000237.3) | c.755T>C p.(Ile252Thr) | P | Lipoprotein lipase deficiency [AR; MIM#238600] \|<br>[High density lipoprotein cholesterol level QTL 11]<br>[AR; MIM#238600] \| Combined hyperlipidemia,<br>familial [AD; MIM#144250] | Yes, partially<sup>c</sup> |
| 17 | F | 31-35 | HLD/HTG |  | c.644G>A p.(Gly215Glu) | P |  | Yes |
| 18 | F | 76-80 | HLD/HTG+Pre-Diabetes |  | c.644G>A p.(Gly215Glu) | P |  | Yes |
| 19 | M | 31-35 | HLD/HTG | APOE<br>(NM_000041.4) | c.478C>T p.(Arg160Cys) | LP | Hyperlipoproteinemia, type III [MIM#617347] \|<br>Coronary artery disease, severe, susceptibility to<br>[MIM#617347] \| Lipoprotein glomerulopathy<br>[MIM#611771] \| Alzheimer disease 2 [AD;<br>MIM#104310] \| ?Alzheimer disease, protection<br>against, due to APOE3-Christchurch [AD;<br>MIM#607822] \| Sea-blue histiocyte disease [AR;<br>MIM#269600] \| ?Macular degeneration, age-related<br>[AD; MIM#603075] | Yes |
| 20 | F | 56-60 | HLD/HTG+T2DM |  | c.460C>A p.(Arg154Ser) | LP |  | Yes |
| 21 | F | 46-50 | HLD/HTG+T2DM+MAFLD/MASH | PPARG<br>(NM_138711.6) | c.362A>G p.(Tyr121Cys) | P | Lipodystrophy, familial partial, type 3 [AD;<br>MIM#604367] \| Insulin resistance, severe, digenic<br>[AD; MIM#604367] \| Diabetes, type 2 [AD;<br>MIM#125853] \| Obesity, severe [AD/AR/Mu;<br>MIM#601665] \| Carotid intimal medial thickness 1<br>[MIM#609338] | Yes |
| 22 | F | 26-30 | HLD/HTG |  | c.362A>G p.(Tyr121Cys) | P |  | Yes |
| 23 | M | 36-40 | HLD/HTG | ABCG5<br>(NM_022436.2) | c.576del p.(Ile193PhefsTer34) | LP | Sitosterolemia 2 [AR; MIM#618666] | Yes<sup>d</sup> |
| 24 | M | 56-60 | HLD/HTG | APOA5 | chr11:g.116652395_116817024<br>del | LP | Hyperchylomicronemia late-onset [AD, MIM#144650]<br>\| Hypertriglyceridemia susceptibility [AD,<br>MIM#145750] | Yes |
| 25 | F | 36-40 | T2DM | GCK<br>(NM_000<br>162.5) | c.787T>C p.(Ser263Pro) | LP | Diabetes mellitus, noninsulin-dependent, late onset<br>[AD; MIM#125853] \| Diabetes mellitus, permanent<br>neonatal 1 [AR; MIM#606176] \| Hyperinsulinemic<br>hypoglycemia, familial, 3 [AD; MIM#602485] \| MODY,<br>type II [AD; MIM#125851] | Yes |
| 26 | F | 26-30 | T2DM | HNF1B<br>(NM_000<br>458.4) | c.544C>T p.(Gln182Ter) | P | Renal cysts and diabetes syndrome [MIM#137920;<br>AD] \| Type 2 diabetes mellitus [MIM#125853; AD] | Yes |
| 27 | M | 36-40 | HLD/HTG+T<br>2DM+MAFL<br>D/MASH | LMNA<sup>b</sup><br>(NM_170<br>707.4) | c.898G>A p.(Asp300Asn) | P | Lipodystrophy, familial partial, type 2 [AD;<br>MIM#151660] \| Cardiomyopathy, dilated, 1A [AD;<br>MIM#115200] \| Charcot-Marie-Tooth disease, type<br>2B1 [AR; MIM#605588] \| Emery-Dreifuss muscular<br>dystrophy 2 [AD; MIM#181350] \| Emery-Dreifuss<br>muscular dystrophy 3 [AR; MIM#616516] \| Heart-hand<br>syndrome, Slovenian type [AD; MIM#610140] \|<br>Hutchinson-Gilford progeria [AD; MIM#176670] \|<br>Malouf syndrome [AD; MIM#212112] \| Mandibuloacral<br>dysplasia [AR; MIM#248370] \| Muscular dystrophy,<br>congenital [AD; MIM#613205] \| Restrictive<br>dermopathy 2 [MIM#619793] | Yes |
| 28 | F | 31-35 | HLD/HTG | MC4R<br>(NM_005<br>912.3) | c.171del p.(Ser58AlafsTer7) | P | Obesity (BMIQ20) [AD/AR; MIM#618406] \| Obesity,<br>resistance to (BMIQ20) [AD/AR; MIM#618406] | Yes<sup>e</sup> |
| 29 | F | 36-40 | T2DM+MAFLD/MASH | NR0B2<br>(NM_021<br>969.3) | c.293_301del<br>p.(Leu98_Ala101delinsPro) | LP | Obesity, mild, early-onset [AD, AR, Mu; MIM#601665] | Yes<sup>e</sup> |
| 30 | F | 26-30 | Pre-Diabetes | SIM1<br>(NM_005<br>068.2) | c.793_794del<br>p.(Leu265ValfsTer101) | LP | ?Obesity with Prader-Willi like phenotype<br>[PMID:23778139, 25351778] | Yes<sup>e</sup> |

Abbreviations: F; female, M; male, HLD/HTG; hyperlipidemia/hypertriglyceridemia, MAFLD; metabolic dysfunction-associated fatty liver disease, MASH; metabolic dysfunction-associated steatohepatitis, T2DM; type 2 diabetes mellitus, LP; likely pathogenic, P; pathogenic, AD; autosomal dominant, AR; autosomal recessive, Mu; Multifactorial.
<sup>a</sup> All variants detected in heterozygous state except for this variant detected in homozygous state.
<sup>b</sup> The P/LP variants here in these genes were also reported as ACMG secondary findings.
<sup>c</sup> This participant has been on statin for a long time although the enrollment diagnosis was T2DM.
<sup>d</sup> This participant also carries a missense VUS, phase undetermined. The participant has been biochemically diagnosed to have Sitosterolemia following the reporting of two variants in ABCG5.
<sup>e</sup> These participants have BMI >30 with the *MC4R* and *SIM1* variant carriers having BMI of 37 each.

A total of 190 VUS identified in 144 participants (25%) were reported. The most commonly reported genes with VUS were *APOB* (n = 38), *ABCC8* (n = 18), *LDLR* (n = 16), *WFS1* (n = 13), *ABCA1* (n = 10), and *ALMS1* (n = 6). The remaining genes were reported one to five times (Supplementary Table 2). While the combination of T2DM and MAFLD/MASH has the highest P/LP pathogenic variant yield, HLD/HTG has the highest among four major isolated indication categories (Supplementary Figure 2).

The proportion of reported VUS was highest among participants self-reporting as Asian (30%) and African American (29%) race/ancestry, while it was 20% among participants self-reporting as White (Supplementary Figure 3).

No variants were reported in about one fourth of participants (n = 133, 24%) while 234 participants (42%) had only variant(s) in the supplementary section. Although participants with P/LP variants tended to be younger, there was no significant difference in age distribution between participants with negative or supplementary results and participants with VUS reported in the primary section (Supplementary Figure 4).

Secondary Findings: Eighteen participants did not consent to receive ACMG secondary finding (SF) results. Among the remaining participants 45 P/LP variants in 15 genes were detected in 40 participants, of which 30 participants had P/LP variants in genes that were not included in the panel gene list (5.5%). Key findings include the c.3920T>A, p.(Ile1307Lys) variant in *APC*, classified as a risk allele for colorectal cancer in participants of Ashkenazi Jewish (AJ) ethnicity, was reported in 9 participants of AJ descent. Variants in *BRCA1/2* and *PALB2* variants were identified in 14 participants of whom 6 were male. *LDLR* and *APOB* variants were identified in 10 participants with one participant carrying two P/LP LDLR variants; these variants were also reported as primary findings due to the HLD/HTG phenotype. Four participants were found to carry more than one P/LP variants in genes included in the ACMG secondary finding list (Supplementary Table 2). In total 30 participants had reportable variants in

## DISCUSSION

This study highlights the genomic findings in participants presenting with complex metabolic diseases and associated comorbidities. A notable proportion of participants (24%) had no reportable variants, while 42% had variants exclusively reported in the supplementary section. Despite the absence of P/LP variants in these participants, the identification of VUS and supplementary findings offers opportunities for reverse phenotyping to uncover potential genotype-phenotype correlations and expand the understanding of genetic contributions to multifactorial diseases.

Interestingly, participants with P/LP variants tended to be younger; however, age distribution differences were not statistically significant when compared to participants with negative, supplementary findings or those with VUS reported in the primary section (Supplementary Figure 4). This observation underscores the complexity of interpreting genomic data in multifactorial metabolic conditions, where clinical utility is often influenced by a combination of genetic, environmental, and lifestyle factors.

The detection of 45 pathogenic/likely pathogenic variants in 15 ACMG secondary finding genes in 7% of participants demonstrate the broader clinical impact of genome sequencing beyond the primary metabolic indications. Notably, the c.3920T>A, p.(Ile1307Lys) variant in *APC*, a known risk allele for colorectal cancer in participants of Ashkenazi Jewish (AJ) descent,^16^ was identified in nine participants, highlighting the significance of ethnicity-specific variant screening. Similarly, actionable variants in *BRCA1/2* and *PALB2* were reported in 14 participants, including six males, emphasizing the role of genomic findings in identifying cancer predisposition syndromes. Importantly, variants in *LDLR* and *APOB* identified in 10 participants further underscore the overlapping roles of genes contributing to both primary indications (e.g., HLD/HTG) and secondary actionable findings.

The identification of participants carrying multiple P/LP variants in ACMG genes (*n* = 4) reflects the value of a comprehensive genomic evaluation in addressing co-existing risks. Such findings necessitate careful clinical follow-up and family-based screening to identify at-risk relatives and inform preventive strategies. For example, the detection of actionable variants enabled several participants to initiate surveillance or personalized interventions, such as the use of setmelanotide for obesity caused by variants in *MC4R*, *POMC*, *PCSK1*, or *LEPR*.^17^

### Clinical and Technical Utility of GS

Our study demonstrates the utility of GS in diagnosing monogenic causes of metabolic diseases. GS offers significant advantages over other next-generation sequencing (NGS) approaches, such as exome sequencing (ES) or targeted gene panels (TGP), due to its ability to:

1. Detect clinically relevant copy number variants (CNVs): For example, a ∼165 Kb deletion in *APOA5* identified in one patient with HLD/HTG contributed to a definitive diagnosis. While CNV detection is possible with other NGS approaches, GS offers genome-wide coverage and higher resolution enabling more accurate identification and improved delineation of CNV breakpoints.
2. Capture variants in deep intronic and untranslated regions: GS enables comprehensive genome-wide coverage, including noncoding regions that may harbor novel or previously unrecognized P/LP variants. For example, in this study, we identified 13 rare variants in genes relevant to the testing indication. While classified as VUS based on the available evidence, several showed high spliceAI scores, making them strong candidates for RNA studies. Such variants may be missed by targeted approaches that only capture previously known variants.
3. Facilitate novel gene discovery and iterative reanalysis: GS enables the identification of previously unrecognized gene-disease relationships due to its comprehensive, genome-wide scope, which surpasses the limits of exome sequencing and targeted panels. While reassessment of VUS over time is possible with all sequencing approaches, GS offers the broadest dataset for iterative reanalysis as new gene-disease associations emerge.

The potential for longitudinal follow-up and integration with large-scale cohorts (e.g., TOPMed, UK Biobank, All of Us) is another key strength of GS.^18–20^ For instance, the association between *KIV2L* repeat numbers and lipoprotein A (Lp(a)) levels^21^ highlights how GS can uncover structural variations contributing to metabolic phenotypes, a focus for future phases of this study. Additionally, GS is also amenable to polygenic risk score calculations in individuals with or without a clinically significant variant in one of the metabolic disease associated genes.

### Challenges in Variant Interpretation

Despite these advantages, our study highlights challenges in variant classification in this disease cohort, particularly for missense variants:

1. In adult-onset multifactorial diseases, classification as likely pathogenic is often more difficult than in pediatric monogenic disorders. This is because multifactorial diseases involve overlapping comorbidities and variable penetrance, reducing the specificity of genotype-phenotype associations. In contrast, pediatric monogenic disorders often exhibit distinct and highly penetrant phenotypes, enabling variant classification based on limited but definitive cases.
2. Population databases such as gnomAD and TOPMed include individuals with common metabolic conditions, making it harder to distinguish pathogenic variants from benign polymorphisms. Age-related penetrance further complicates interpretation by introducing uncertainty about whether individuals with variants will eventually manifest disease.

These factors likely contribute to the relatively low diagnostic yield (6%) in our study. However, additional contributors include the limited 90-gene panel used for primary interpretation and the inclusion of individuals with early or subclinical phenotypes (e.g., prediabetes). Improvements in variant interpretation will also require integrated approaches using machine learning, transcriptomics, proteomics, and functional assays such as CRISPR-based mutagenesis screenings.

### Study Limitations and Future Directions

This study has several limitations. First, our analysis was restricted to a pre-selected 90-gene panel, which may exclude significant variants in genes not currently associated with metabolic diseases. Future unbiased genome-wide analyses may identify novel genes and risk variants contributing to metabolic phenotypes. Second, the cohort was enriched for participants with HLD/HTG and obesity, as referrals were primarily from specialty clinics, and it also included individuals with prediabetes in whom genetic causality may be more difficult to establish (Figure 1). Thus, our findings may not be generalizable to broader or more diverse populations. Finally, further studies, including functional validation of VUS, longitudinal follow-up, and reverse phenotyping, are needed to enhance diagnostic yield and clinical interpretation.

## Conclusion

In conclusion, we report a 6% diagnostic yield for monogenic causes of multifactorial metabolic diseases through the use of GS in this cohort of participants with metabolic diseases. While the current yield is modest, GS has already provided actionable findings that influenced clinical management and surveillance. As genomic technologies evolve, continued efforts in functional studies, reanalysis of existing data, and large-scale population studies will enhance our ability to identify and classify genetic variants. Future studies should also assess the cost-effectiveness of GS in metabolic disease clinics and explore its role in early diagnosis, prevention, and targeted therapy for individuals with complex metabolic conditions.

## SUPPLEMENTARY INFORMATION

Supplementary information contains Supplementary Tables 1-2 and Supplementary Figures 1-4.

## DATA AVAILABILITY

The authors will make relevant phenotypic, biochemical, and molecular genetics data available for the purposes of verifying or contextualizing the conclusions they have drawn in this publication. Any disclosure will be constrained by the need to protect the privacy of respondents. Transcripts of clinical consultations cannot be made available; however, the authors will consider requests to supply relevant, unpublished additional segments of consultation data for the purposes of verifying or contextualizing the conclusions. Phenotypic and biochemical data requests should be sent to the corresponding author M.D.G. and molecular genetics data to the corresponding author V.J. with a description of the reason for the request and the qualifications of those requesting the data. Individual variants reported in this study have been deposited in ClinVar by the New York Genome Center.

## ACKNOWLEDGEMENTS

The authors would like to thank all the study participants for their time and insights into their experiences.

## Author Contributions

Conceptualization and Methodology: V.J., M.D.G.; Data curation: V.O., A.U.T, A.T.W., S.G., A.A., A.L.W., S.Y.K., S.T.S, S.P., P.K., C.N., V.J.; Visualization: V.O.; Formal analysis: V.O.; Writing-original draft: V.O., A.H., J.N.F; V.J., M.D.G.; Writing-review and editing: V.O., A.H., J.N.F, M.A.H., S.L.S., K.C., J.M., F.D., S.K., M.Y., G.D., O.B.M., C.A., L.C.H., E.W., L.G., A.K., J.T., A.U.R., A.T.W., S.G., A.A., A.L.W., S.Y.K., S.T.S., S.P., C.N., S.C., R.S., T.Y.M., M.J.R., L.C.A., O.E., M.S.U., J.M.P., V.J., M.D.G; Software: A.A., P.K.; Resources: A.H., J.N.F., M.A.H., S.L.S., K.C., J.M., F.D., S.K., M.Y., G.D., O.B.M., C.A., L.C.H., E.W., L.G., A.K., J.T., S.C., R.S., T.Y.M., M.J.R., L.C.A., O.E., M.S.U., J.M.P., V.J., M.D.G.; Project Administration: M.A.H., R.S.; Funding Acquisition: V.J., M.D.G.; Supervision: V.J., M.D.G.

## Ethics Declaration

Ethical approval for this research was obtained from the Institutional Review Board of Weill Cornell Medical College. Informed consent to participate in this study was obtained from all participants.

