## Supplemental Figure for "Genome Sequencing Identifies Monogenic Causes in Adults with Metabolic Diseases"

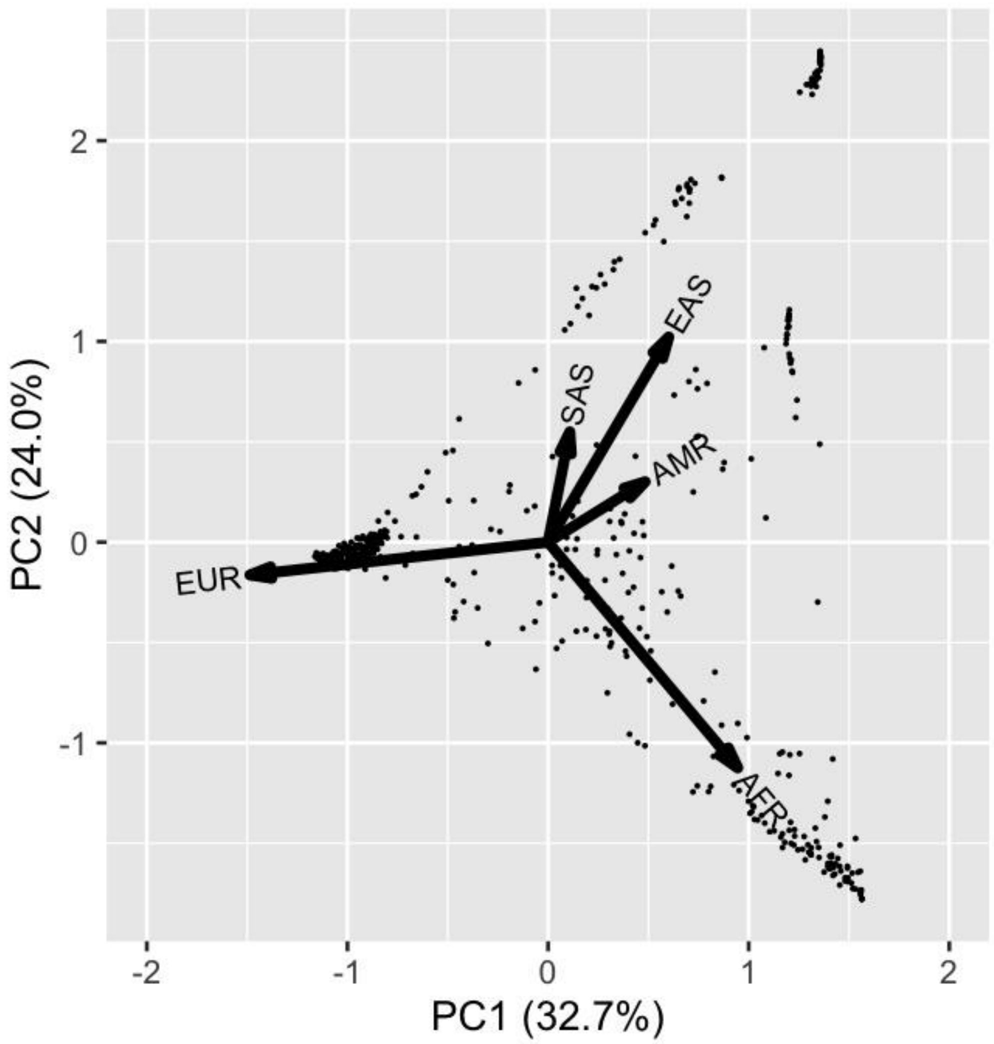


**Figure S1**. Principal component analysis of ancestries among participants.

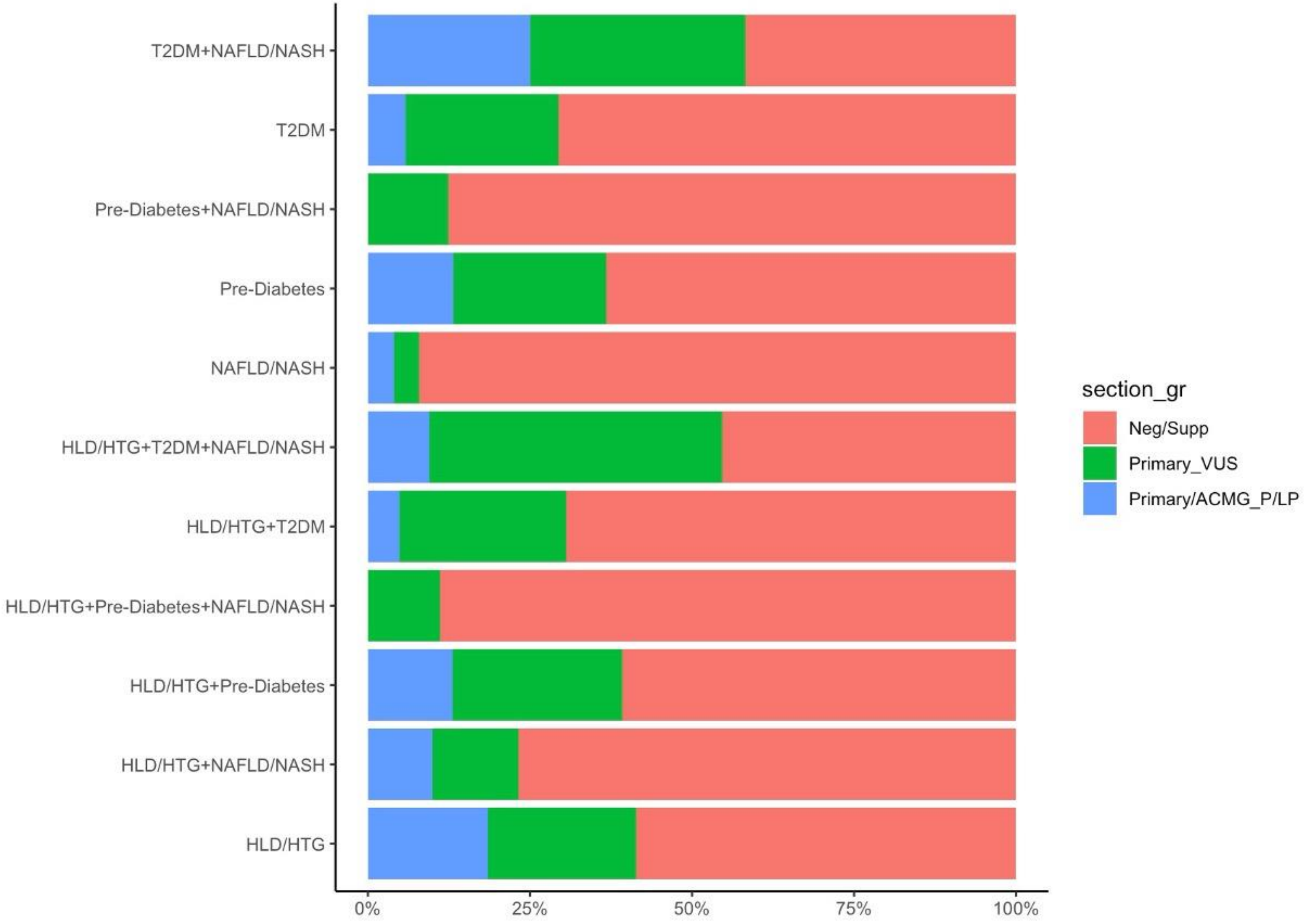


**Figure S2**. Variant detection percentages based on indications.

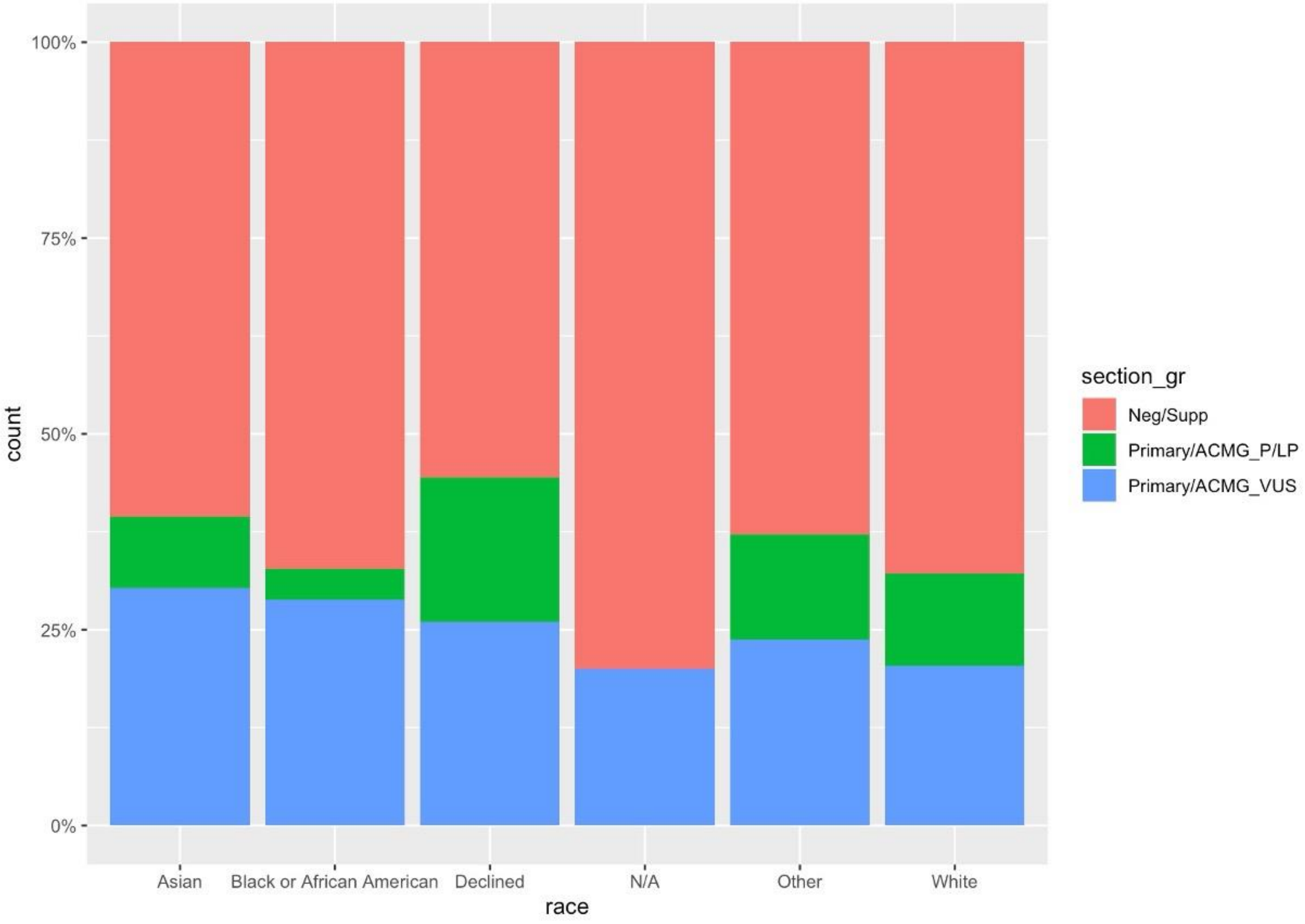


**Figure S3**. Variant detection percentages based on self reported ancestry.

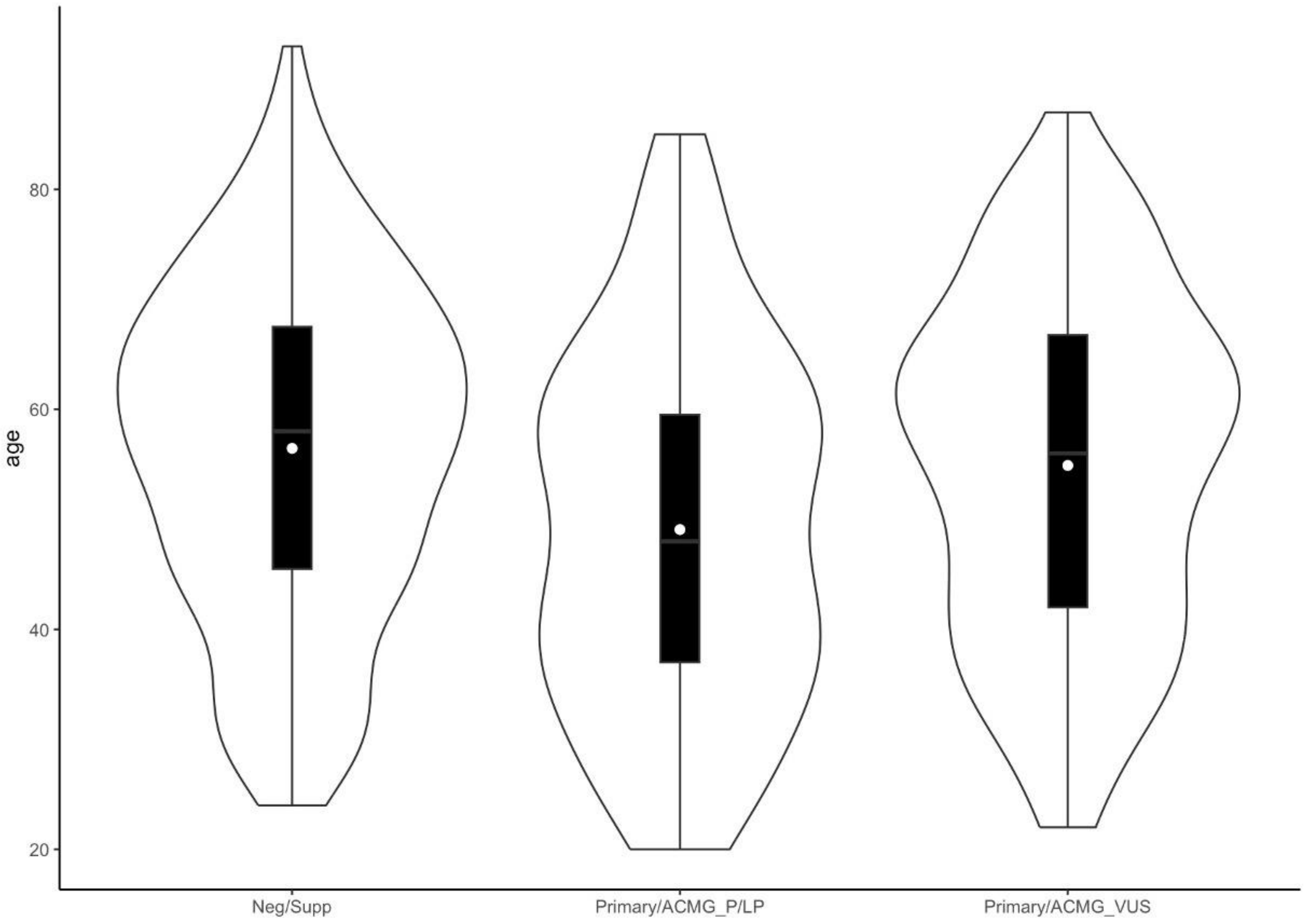


**Figure S4**. Variant detection percentages based on age.
